# Finding the groove in neural space

**DOI:** 10.64898/2026.02.26.26347169

**Authors:** Rashi Bhatt, Drew E. Sheets, Patrick M. Jordan, John E. Downey, Hugo Merchant, Charles M. Greenspon

## Abstract

The neural signature of rhythm and tempo remains difficult to study in both humans and non-human primates. Here we recorded from the motor cortex of human participants implanted with intracortical microelectrode arrays while they performed a series of rhythmic tapping tasks. We found that rhythmic tapping elicited low-dimensional rotational neural dynamics whose radii varied in a tempo-dependent manner and axes related to kinematic properties. Moreover, we observed a spectrum of kinematic and neural behavior as participants shifted from low tempo punctuated taps to high tempo smoother, continuous taps. Surprisingly, we observed that tactile feedback strengthened the rotational dynamics despite reduced kinematic range. Moreover, while tempo preparation did not produce dynamics of their own, motor cortex encoded it in an orthogonal dimension. Finally, we found that switching tempos was achieved with smooth neural transitions that could only be separated in higher dimensions. These results show that motor cortex directly encodes a multitude of rhythm related features.

## Introduction

Music cognition is phenomenon shared across human cultures^1,2^. Specifically, humans are able to extract salient periodic events – known as beats or pulses – from songs and then align their movements for music and dance^3,4^. This requires coordination of many neural structures as one must attend to the music, integrate and analyze its temporal structure, and then create an appropriate motor plan. These motor plans are often interrogated through the analysis of tapping movements which are entrained to an internally generated beat and typically precede the external cues by several milliseconds^5,6^. When asked to tap at a comfortable tempo, people reliably produce beats with intervals between 500 and 600 ms (approximately 120 beats per minute, BPM) which is known as the spontaneous motor tempo^7^. Interestingly, this spontaneous tempo is consistent with the preferred tempo of music across human cultures^8^. Functional imaging and extracranial recordings in humans have implicated many regions in beat generation including medial motor and premotor areas, the basal ganglia, and the cerebellum^9–14^.

Recent neurophysiological studies in monkeys indicate that the neural representation during rhythmic tapping to visual metronomes depends on the neural population dynamics in medial premotor cortex (MPC)^15^. A key property of MPC neurons is the relative representation of beat timing: they are progressively recruited in rapid succession producing neural sequences that adaptively fill the beat duration depending on the tempo to provide a linear representation of interval evolution^15,16^. Moreover, projecting MPC activity into a low-dimensional space reveals a distinct rhythm representation^17,18^ First, they have oscillatory dynamics that form a repeating loop for every produced interval. Second, they converge to a single point in state space for each tap. Third, the periodic trajectories increase in amplitude and decrease in speed to encode tempo^17,18^. Finally, experiments using auditory and visual metronomes reveal this representation to be amodal^18^.

Here we investigated the neural signatures of rhythm in the hand representation of motor cortex in two human participants with spinal cord injuries implanted with intracortical microelectrode arrays. We assess the dynamics associated with spontaneous and entrained tapping, the representation of continuously varying tempo, the interaction between kinematics and sensory feedback, and the dimensionality required to flexibly change between tempos.

## Results

### Internally generated rhythmic movements evoke rotational dynamics in motor cortex

We first sought to determine how rhythmic movements were represented in the absence of any externally provided cues as NHP research can suffer from overtraining. Consequently, we asked one participant (C1) to first tap at a “comfortable” rate with the expectation that it would correspond to their spontaneous motor tempo – and gave no further instructions. During the ∼30 seconds of tapping, we recorded neural activity from both sensory and motor cortices and kinematics from a camera viewing their hand and a capacitive touch sensor that they tapped (**Figure 1A**). Then, we asked C1 to tap at either a “slow” or “fast” pace so that we could compare the neural activity. He produced 3 distinct tempos with consistent inter-tap intervals (Figure 1B, **Supplementary Figure 1A**, 1.2-0.2 seconds/60-300 beats per minute) and produced smooth sinusoidal kinematics (**Supplementary Figure 1B**), during which many neurons fired cyclically, particularly during the slow tempo (**Figure 1C**). Applying dimensionality reduction techniques, we observed continuous rotational dynamics in the first three principal components of the neural data (**Figure 1D, Supplementary Figure 1C**), consistent with previous NHP research^17,19,20^. Similarly, the tempo at which the participant was tapping was seemingly represented by the diameter of the rotations (**Figure 1E**), with minimal separation of the tempos along the third axis. Surprisingly, the neural trajectories appeared to converge at the apex of the tap sequence and not at the tap event itself, similar to what is observed with RNNs trained on a similar task^21^. Finally, we sought to determine if these dynamics were present in both motor and sensory cortices. Given the apparent rotations, we used jPCA^22^ to find the optimal rotational plane within the neural activity of each region. We observed that the rotational dynamics were much more prominent in motor cortex, while sensory cortex activity only varied along a single dimension (**Figure 1F**, **Supplementary Figure 1D**). This shows that the rotational dynamics previously observed in NHPs^17,23^ are not a result of overtraining but instead an instrinsic property of cyclical motor behavior.

**Figure 1.**
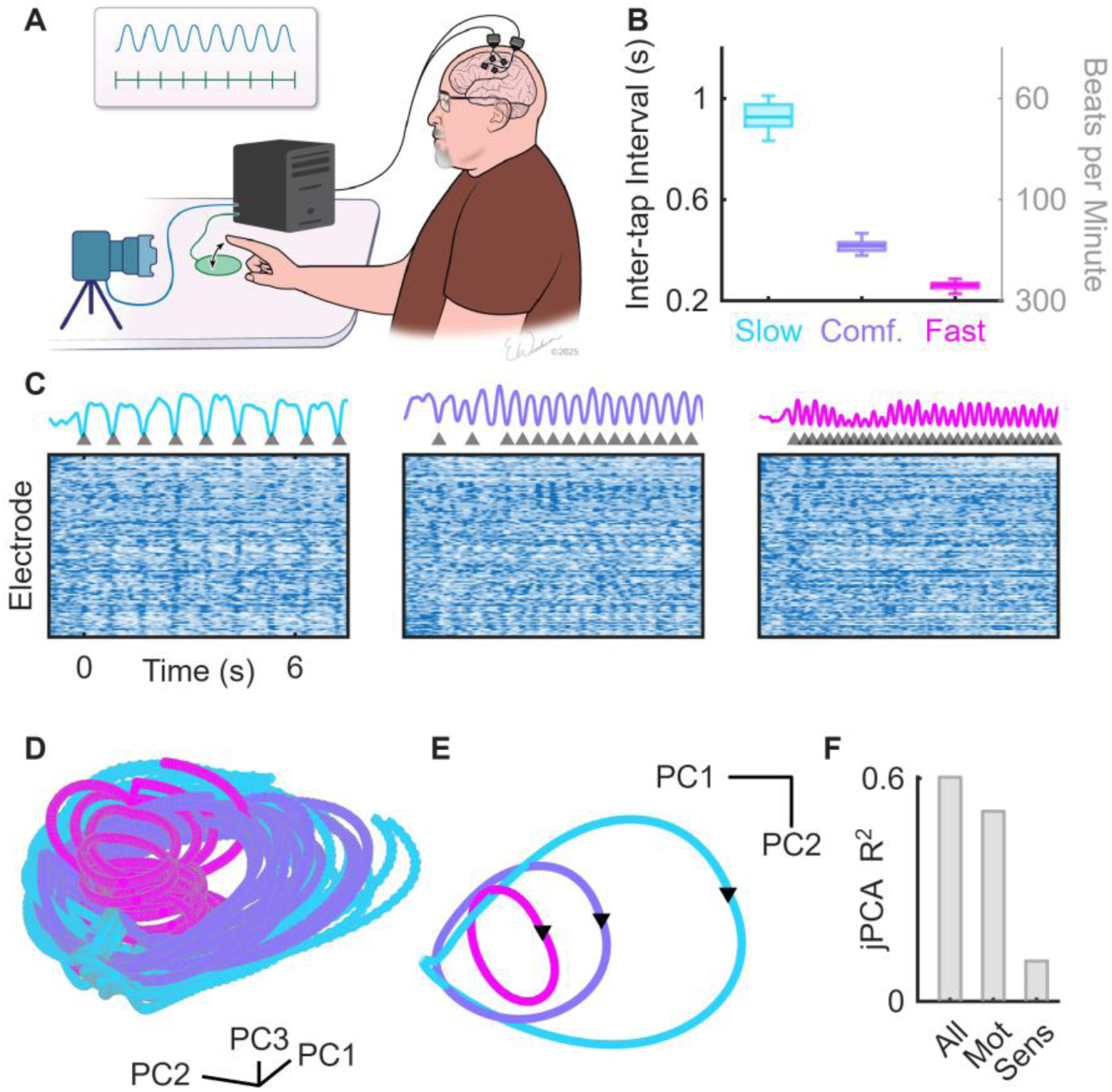
Unconstrained rhythmic tapping behavior produces neural rotations. **A)** Experimental setup. Participants implanted with Blackrock NeuroPort arrays in motor and sensory cortex tapped a capacitive sensor while a camera recorded finger position. **B)** Inter-tap intervals from C1 when instructed to tap ‘slowly’, ‘at a comfortable speed’, or ‘fast’ in both seconds (left) and beats per minute (right). **C)** Kinematic and neural data. Lines show finger position, triangular markers indicate tap events, and the heatmap shows Z-scored neural activity where white is low and blue is high. **D)** Low dimensional projections of the neural data. **E)** Tap-averaged trajectories for each tempo. Black markers indicate tap event as shown in **B**. **F)** Explained variance of the first jPCA rotational plane fit to different groups of electrodes. Data from C1.

### Rhythm dependent dynamics reflect underlying kinematics

Having established the presence of rotational dynamics during rhythmic tapping, we wanted to better examine the relationship between tapping speed, its entrainment, and the resulting neural dynamics. Consequently, we cued two participants (C1 and C2) to tap at 7 equally spaced tempos between 30 and 210 beats per minute via auditory tones. Before each trial, a distinct audio cue was played to indicate that the tap cues would begin in ∼3 seconds.

Examining the kinematics (**Figure 2A**), the participant tended to miss the first 1-2 tones while adjusting to the tempo and reached a stable tapping latency by the 5^th^ tap (**Supplementary Figure 2A,B**), and exhibited classical negative lag-1 cross-correlations consistent with error correction^6,20^ (Pearson correlation: r = −0.239, p < 0.01, **Supplementary Figure 2C**). We therefore defined the initial phase (∼5 taps) as “entrainment” and the subsequent phase as “maintenance”. Additionally, we observed that in the slower tempos the participant tended to exhibit punctuated, non-sinusoidal, kinematics in which the inter-tap interval contained long dwell periods with an elevated hand followed by brief taps near the tone (1-way ANOVA: cosine tuning, F_[6,69]_=81, p<0.01; dwell time, F_[6,69]_=71, p<0.01, **Supplementary Figure 2D,E**), similar to NHPs^20,24^, and were slightly larger than taps at the higher tempos (F_[6,3610]_=215, p<0.01, **Supplementary Figure 2F**).

**Figure 2.**
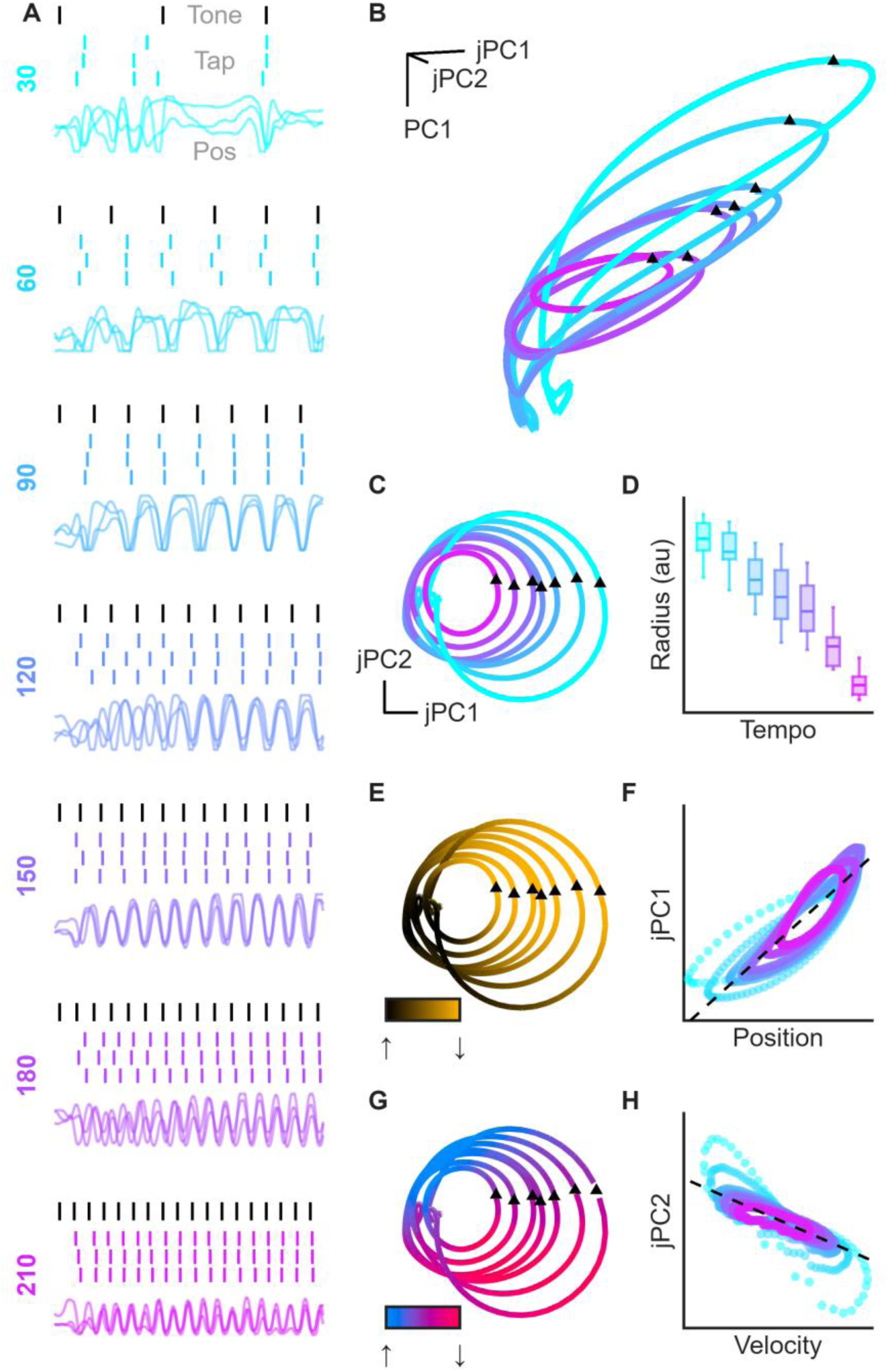
Neural dynamics across discrete tempos. **A)** Example kinematics. Black ticks indicate tone times, colored ticks indicate the time at which the participant tapped, and colored traces indicated the position of the fingertip over time. Y-labels indicate target tempo. **B)** Tap-averaged trajectories during the entrained period colored by tempo. Triangles indicate tap events. **C)** Rotational plane view of the data shown in **B**. **D)** Radius distribution for each tempo. **E)** Same view as **B** but colored by the normalized finger position for each tempo. **F)** Relationship between tap-averaged position and jPC1 projection for each tempo. **G)** Same view as **B** but colored by velocity for each tempo. **H)** Relationship between tap-averaged velocity and jPC2 projection for each tempo.. Data from C1.

Building on the previous experiment, we used a combination of jPCA and PCA (see Methods: Dimensionality reduction) to identify the 3-dimensional space that best explained the neural activity across tempos. Plotting the tap-averaged trajectories during the maintenance phase (**Figure 2B, Supplementary Video 1**), we observed rotational dynamics across all tempos that overlapped in the 3^rd^ dimension. Surprisingly, low tempo trajectories also exhibited an excursion along the residual principal component during the dwell phase that was not present at the higher tempos. Next, we sought to examine if there was a change in the dynamics during the transition between entrainment and maintenance phases. We computed the slope of the distances between the centroids of the first 5 taps and the mean centroid of the maintenance period. This revealed consistently negative values, suggesting a convergence of the rotations to the stable point (Sign Test, s=19/70, p < 0.01, **Supplementary Figure 2G**). Additionally, there was an effect of tempo on the convergence rate (1-way ANOVA: F_[6,69]_=3.1, p=0.01) in which the 120 BPM tempo converged the quickest, perhaps due to the fact that it was closest to spontaneous motor tempo^7^.

Given that the rotational dynamics during the internally generated taps seemingly showed tempo-dependent radii (**Figure 1E**), we sought to re-examine this effect. Computing the radius in the rotational plane (**Figure 2C,D**), we found a linear tempo-dependent effect where radius decreased with tempo (1-way ANOVA: F_[6,69]_=65, p<0.01). Moreover, trends between tempo and trajectory length and speed were also observed (**Supplementary Figure 2H,I**) while trajectory length, speed, and radius were all intertwined (**Supplementary Figure 2J-L**) implying that neural dynamics are constrained by the kinematics. As tempo is explicitly related to kinematics, we investigated the relationship between the kinematic range and the trajectory radius, finding a positive correlation (**Supplementary Figure 2M**). Furthermore, we examined the relationship between the neural trajectories and kinematic properties. We found that finger position varied along the first jPC (**Figure 2E,F**, r=0.90, p<0.01) while finger velocity varied along the second (**Figure 2G,H**, r=-0.82, p<0.01). Finally, we examined the rotational strength across tempos and found it to be consistently counterclockwise except during the dwell phase for slower tempos (**Supplementary Figure 2N,O**). Participant C2 replicated these findings, while the nature of his injury resulted in less stable kinematics (**Supplementary Figure 3A-D**), the relationship between neural dynamics and kinematics held across tempos (**Supplementary Figure 3E-J**). These results demonstrate that both position and velocity are simultaneously encoded in motor cortex and implies that tempo-dependent differences are driven by kinematic differences.

### Finding a continuous tempo axis in neural space

With this foundation in place, we next sought to determine if we could find an independent axis that encoded tempo as the first residual PC in the single tempo task could not separate the conditions (**Figure 2B**). We thus constructed a task in which participant C1 either slowly ramped up from 30-210 BPM or the reverse across ⁓3 minutes (**Figure 3A**). The participant was able to closely follow both ramps (Pearson correlation: r=0.94, p<0.01), though there were occasional missed or double taps (**Supplementary Figure 4A**). Across tempos we observed similar shifts from punctuated to smooth continuous taps that coincided not with substantial changes in kinematic range but instead with changes in finger velocity (**Supplementary Figure 4B-E**), though range and velocity were themselves correlated (**Supplementary Figure 4F**). As in the single tempo experiment, correlations were observed between trajectory radius and other metrics including trajectory speed, kinematic range, and instantaneous tempo (**Supplementary Figure 4G-J**).

**Figure 3.**
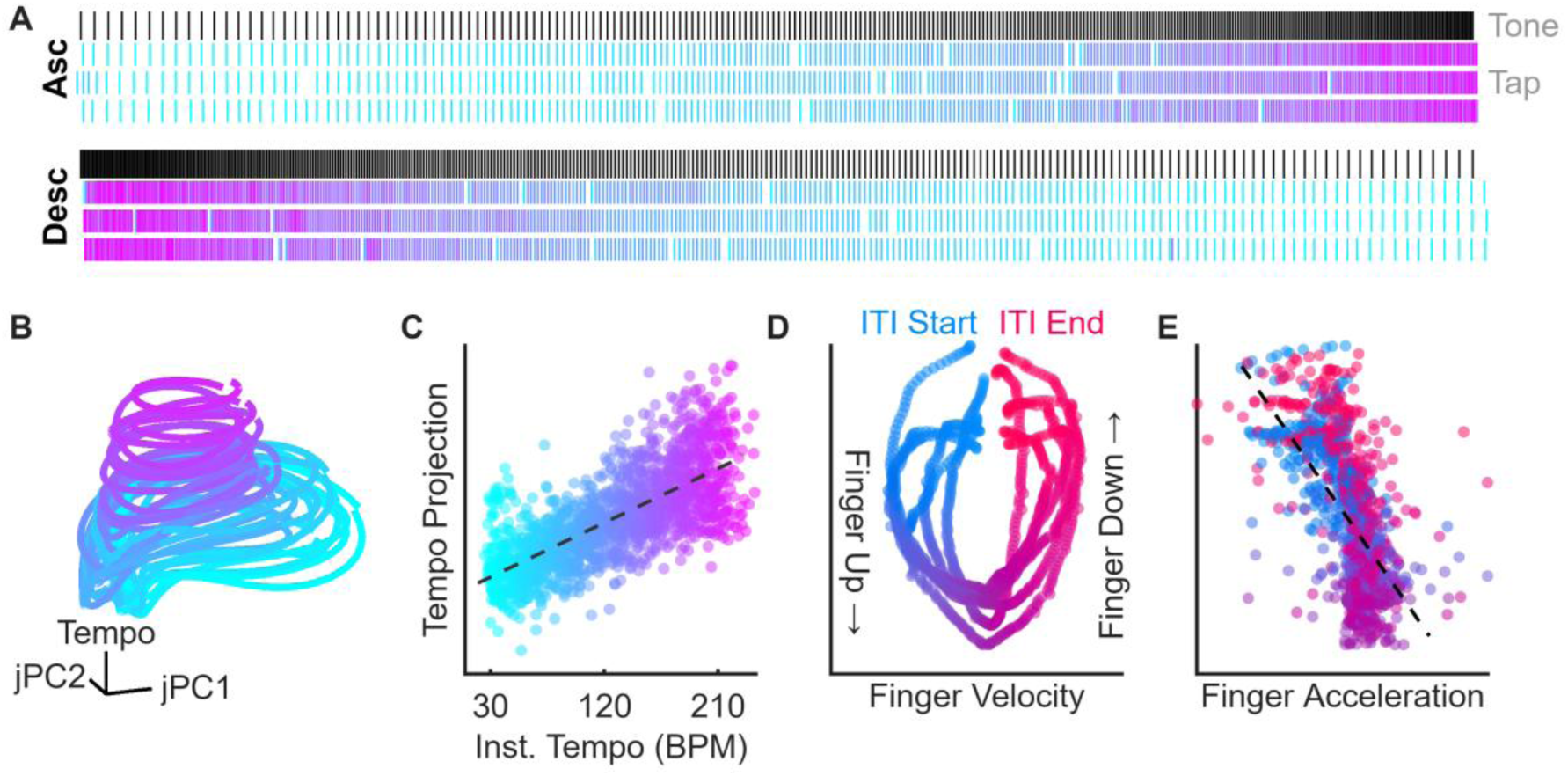
Finger velocity and acceleration are encoded along a tempo axis. **A)** Tone and tap times for ascending and descending trials. Tap markers are colored by their ‘instantaneous’ tempo. **B)** Neural data projected onto the rotational plane and the tempo axis. Each trajectory represents the average trajectory of 10 taps for visualization. **C)** Relationship between the instantaneous tempo for each tap and the tap averaged tempo axis projection. **D)** Relationship between the tap-averaged finger velocity or **E)** acceleration and tempo projection. Points are colored by progression through tap sequence.

To identify the putative tempo axis, we applied a combination of jPCA, PCA, and multiple linear regression to find both the optimal rotational plane and the combination of residual PCs that best predicted tempo. This axis separated the trajectories and correlated with both the instantaneous tempo and finger velocity (**Figure 3B,C**, r = 0.746; **Supplementary Figure 4K,L**). Notably, this axis was able to accurately predict instantaneous tempo better than any of the individual residual PC (r = −0.230 to 0.461). Given the apparent relationship between the tempo axis and finger velocity, we sought to examine if the axis was sensitive to changes in velocity within the tap cycle. Taking the average tempo axis score and velocity at each point across all taps in each trial, we found that there were small but consistent non-linear within-tap differences where neutral velocities during the dwell period were associated with low tempo axis values and either positive or negative velocities were associated with high tempo axis values (**Figure 3D**). Converting to acceleration, we found a negative relationship where higher acceleration values surprisingly had lower tempo axis values (r=-0.64, p<0.01, **Figure 3E**). This reveals that all kinematic variables necessary to produce rhythmic tapping - position, velocity, and acceleration - are both present and orthogonally encoded in motor cortex.

### Tactile feedback enhances motor cortex dynamics

Two consistent features of the previous tasks were that the participants attempted to align the physical tap event with the tones. This likely resulted in sensory feedback driving neural activity in motor cortex which may influence the neural dynamics, even if the neural activity in tactile sensory cortex wasn’t inherently rotational (**Supplementary Figure 1D**). As it is common for spontaneous rhythmic behavior to include the tapping of one’s foot or fingers on a surface, we questioned whether this tactile feedback was critical both for consistent behavior and neural dynamics. To achieve this, we asked participant C1 to either tap the capacitive plate (as with previous experiments) – Feedback (FB) - or perform the taps “in air” in which they never contacted the plate or any other surface – No Feedback (NFB) -at 3 different tempos (**Figure 4A**, 30-150 BPM).

**Figure 4.**
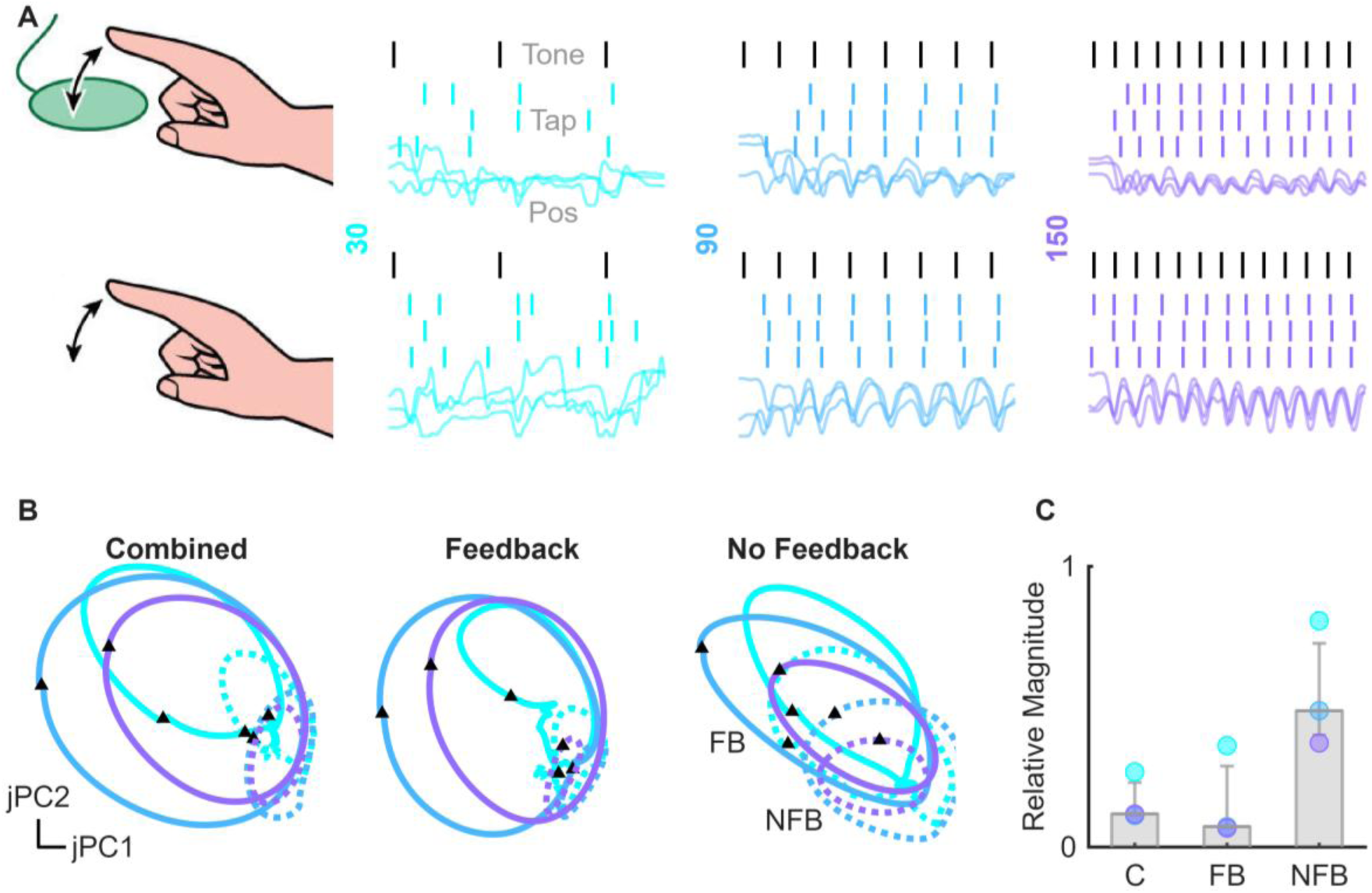
Air tapping task and resultant neural dynamics. **A)** Task illustration in which the participant tapped the capacitive plate or tapped ‘air’ with example trials for each condition. Black ticks indicate tones, colored ticks indicate taps, lines indicate finger position. **B)** Neural trajectories projected into the rotational planes computed on all trials, just the Feedback (FB) trials, or just the No Feedback (NFB) trials. Solid lines indicate Feedback trials, dotted lines No Feedback trials. Triangle markers indicate tap events. **C)** Ratio of NFB variance over FB variance for each tempo projected onto each of the rotational planes.

First, we questioned to what extent the kinematics differed as a result of removing tactile feedback. We found that there was a significant increase in the kinematic range (2-way ANOVA: Tempo, F_[2,29]_=22.84, p<0.01; Task Type, F_[1,29]_=126.77, p<0.01, Tempo X Task Type: F_[2,29]_=10.44, p<0.01, **Supplementary Figure 5A**). Second, despite the lack of a clear alignment event, there was no change in the stability of the taps (Tempo, F_[2,29]_=5.04, p=0.015; Task Type, F_[1,29]_=5.06, p=0.624, Tempo X Task Type: F_[2,29]_=10.07, p<0.01, **Supplementary Figure 5B**). Considering the neural data, the exaggerated kinematics led us to expect similarly exaggerated dynamics, however we instead saw that the rotations were smaller regardless of which data the rotational plane was fit to (**Figure 4B,C**). This is particularly surprising given the previously observed relationships between kinematic range and trajectory radius (**Supplementary Figure 2M**, **Supplementary Figure 3H**, **Supplementary Figure 4I**).

We reassessed the relationship between kinematic range and trajectory radius (**Supplementary Figure 5C**) and found that while correlations were moderately positive within each condition (r=0.139 and 0.252 for FB and NFB, respectively, p<0.01 for both), differences between conditions were substantial enough to instead produce a negative correlation when considered together (r=-0.249, p<0.01). Reasoning that the smaller radii in the NFB condition might be attributed to a reduction in neural firing rates, we quantified neural activity between conditions for the sensory and motor arrays and found that the firing rates were indeed lower in both regions, but that this reduction was greater on the sensory arrays (**Supplementary Figure 5D**). Critically, given that the participant’s sensory arrays are implanted in Brodmann’s Area 1, this is consistent with a lack of tactile inputs, while the exaggerated kinematics in the NFB condition would likely evoke more activity in the proprioceptive Brodmann’s 3a. While the observed rotational dynamics are not wholly dependent upon tactile feedback, these results show that their magnitude is heavily dependent upon non-rotational tactile feedback.

### Tempo preparation is orthogonal to executed tapping and does not produce rotational dynamics

Rhythmic movements often happen spontaneously in the presence of rhythmic cues. However, unlike in the previous tasks, people take time to assess the tempo and smoothly join the tempo. Consequently, we examined if a preparatory period with auditory cues but without tapping would produce neural dynamics. Participant C1 was presented with trials at 3 tempos while a visual cue indicated whether to tap. On half of the trials tapping he was cued to begin with the first auditory tone while on the other half he was cued to begin tapping after 10 seconds of auditory cues during which they were asked to prepare for the tempo without imagining tapping at the tempo. Moreover, we instructed the participant to continue tapping at the same tempo for a further 10 seconds after the auditory tones completed to assess their ability to maintain the tempo in the absence of a cue (**Supplementary Figure 6A**) which we refer to as preparation, execution, and continuation phases.

Comparing the inter-tap intervals at the start of the execution period, we found no effect of either tempo, preparation, or an interaction between the two (2-way ANOVA, p>0.05 for all factors, **Supplementary Figure 6B,C**). Next, examining the inter-tap intervals at the end of both the execution period and the continuation period, again the participant was able to maintain the tempo almost perfectly, though there was a weak effect of tempo and an interaction between tempo and period (2-way ANOVA: Tempo, F_[2,179]_=6.88, p<0.01, Period, F_[1,179]_=0.03, p=0.85, Tempo X Period, F_[2,179]_=3.68, p=0.03, **Supplementary Figure 6D-F**). Given this lack of variability in the execution and continuation periods, we did not attempt to compare the corresponding neural trajectories and considered continuation taps executed taps for further analyses.

Reaching literature in NHPs has shown that a movement-null subspaces exists that explain movement preparation^25^. Consequently, we hypothesized that tap preparation might result in dynamical activity orthogonal to the previously observed rotations. We used jPCA to find the optimal rotational plane during either the preparation or execution phases. When projecting into the execution space (**Supplementary Figure 6G**), we observed minimal preparation related activity. Similarly, when we fit a rotational plane to the preparation period, we observed only weakly rotational dynamics (jPCA R^2^=0.072, **Supplementary Figure 6H**). Next, we used PCA across both periods to find a shared space (**Supplementary Figure 6I**). As expected, rotational activity was present during the executed taps, but more interestingly, there was moderate separation in the neural activity during the preparation period. Indeed, using linear discriminant analysis on the average PCA scores in any of the PCA subspaces, we were able to classify which tempo the participant was preparing for (**Supplementary Figure 6J**). This suggests that despite the lack of dynamics, tempo preparation may still be encoded in motor cortex.

### Mixed tempo tapping demonstrates flexible switching between nested neural trajectories

A hallmark of music and rhythm is that the temporal structure is complex, not constant or monotonically changing as the previous tasks used. Consequently, we sought to examine neural dynamics as we progress through known tempo sequences. We thus constructed 2 types of trial structure in which the participants would alternate between 2 beats. In the first type (type A, **Figure 5A**), the trials were composed of a singular “slow beat” at the primary tempo and then several “fast beats” at the secondary tempo. In the second type (type B, **Figure 5B**), each bout of fast beats was flanked by a slow beat. These can be envisioned as a repeating beat with an elongated pause (1,2,3, pause, 1,2,3) and a gallop rhythm (1-1,2,3, 1-1,2,3), respectively. As humans naturally gravitate towards rhythms with integer ratios^26^, the majority of the secondary tempos were integer ratios of one another (**Figure 5C**, **Supplementary Figure 7**).

**Figure 5.**
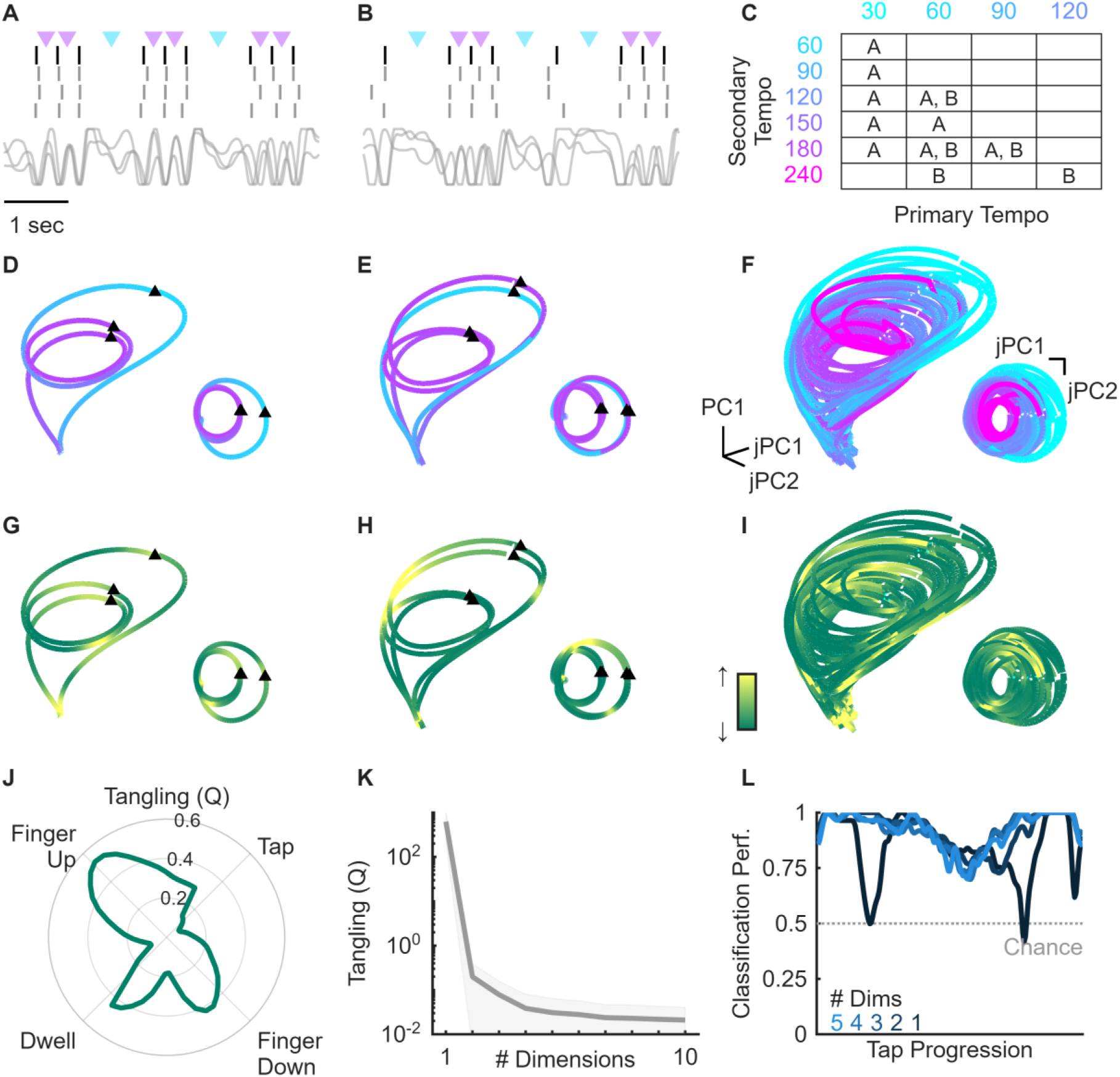
Mixed tempo task and trajectories. Example trial tones, taps, and kinematics for the two types of mixed tempo trials. **A)** Example ‘type A’ trial in which the primary slow tempo (60 BPM) was encoded in the pause between bursts of secondary tempo (120 BPM). **B)** Example ‘type B’ trial in which secondary (120 BPM) tempo was flanked by instances of the primary tempo (60 BPM). **C)** All combinations of type A and B trials that were tested. **D-F**) Sequence averaged trajectories for the trials corresponding to panel A, B, or all trials, respectively. **G-I)** Trajectories from **D-F** color coded by normalized 3D tangling. **J)** Condition average 3D tangling as a function of 2D neural phase. **K)** Tangling across conditions as a function of the number of dimensions the neural data is projected into. **L)** Primary/secondary tempo classification performance as a function of the number of dimensions.

As described previously, we identified a low-dimensional space using jPCA and PCA and projected sequence-averaged data into it (**Figure 5D-F**). Strikingly, we observe that the trajectory smoothly transitions between the large and small radii that varied with the tempo of each tap. Moreover, much like the single tempo experiment, this effect is conserved across the entire sequence such that the first point of the sequence is directly adjacent to the last point of the sequence. Notably, however, while the radii are seemingly distinct between the primary and secondary taps of each condition (paired t-test: p<0.01), the two radii are not aligned at the origin but instead along one edge of each rotation, a feature made more apparent when observed in 2-dimensional space (**Figure 5D-F insets**). The consequence of this is that there is ambiguity in the neural dynamics at these locations.

To visualize this ambiguity, we first computed 2D vector fields for each condition (**Supplementary Figure 8A-C**) where we consistently observed smaller vectors where trajectories overlapped. Then, to quantify this effect, we computed the degree to which there was tangling within each condition (**Figure 5G-I**), a metric that allows estimation of trajectory predictability in higher dimensions^27^. We observed in 3D, consistent with the vector field, that high tangling was localized to three distinct phases of the tap (**Figure 5J**): the dwell period, and the rising and falling phase of the finger where the transition between tempos occurs. Given the stable behavioral performance, this implies that there may be greater separation of the trajectories in higher dimensions. Consequently, we computed tangling in ascending cumulative dimensions (**Figure 5K**) and observed that tangling reduced with increasing dimensions and seemingly plateaued by the 5^th^ dimension. To confirm this, we attempted to classify the tempo within (**Figure 5L**) and across (**Supplementary Figure 8D**) conditions along the tap sequence with increasing dimensions. Performance qualitatively matched the tangling result, where clear periods of low classification are observed using 1-2 dimensions, but that performance plateaued by the 3^rd^ dimension. Critically, we repeated both classifications using individual dimensions and found that performance was consistently worse than using cumulative dimensions, revealing that no one dimension was sufficient to separate the tap tempo across the tap cycle (**Supplementary Figure 8E-F**). These results show that 5 dimensions are sufficient to enable smooth untangled trajectories while alternating tempos.

### Neural geometry is shared across tasks

For each task we computed a separate space using jPCA and PCA before performing the analyses and found conserved structures. These similarities imply both that shared neural mechanisms may persist across tasks and that the observed relationships between neural data and kinematics should hold across tasks. Consequently, we sought to determine how similar the computed neural spaces were between tasks and if we could reliably decode finger position across tasks. Crucially, as speed, tempo, and dwell period can all be derived from the finger position, this means that decoding finger position is representative of the participants behavior. First, we projected each task’s neural data into each task space (jPC1,2 + PCs1-3, **Figure 6A**, **Supplementary Figure 9A**). While there was some similarity between the single tempo and ramp projections, the single tempo trajectories projected into the ramp space were unrecognizable. To quantify this, we computed the cross-projection alignment index^28^ between each task (**Figure 6B**) and found that, indeed, there was a high degree of similarity between the single and syncopated tempo tasks, but a great degree of misalignment between ramp and each task. When computing the absolute angle between the spaces, we found that they were only partially aligned, suggesting that some neurons contributed quite differently to each task (**Supplementary Figure 9B**).

**Figure 6.**
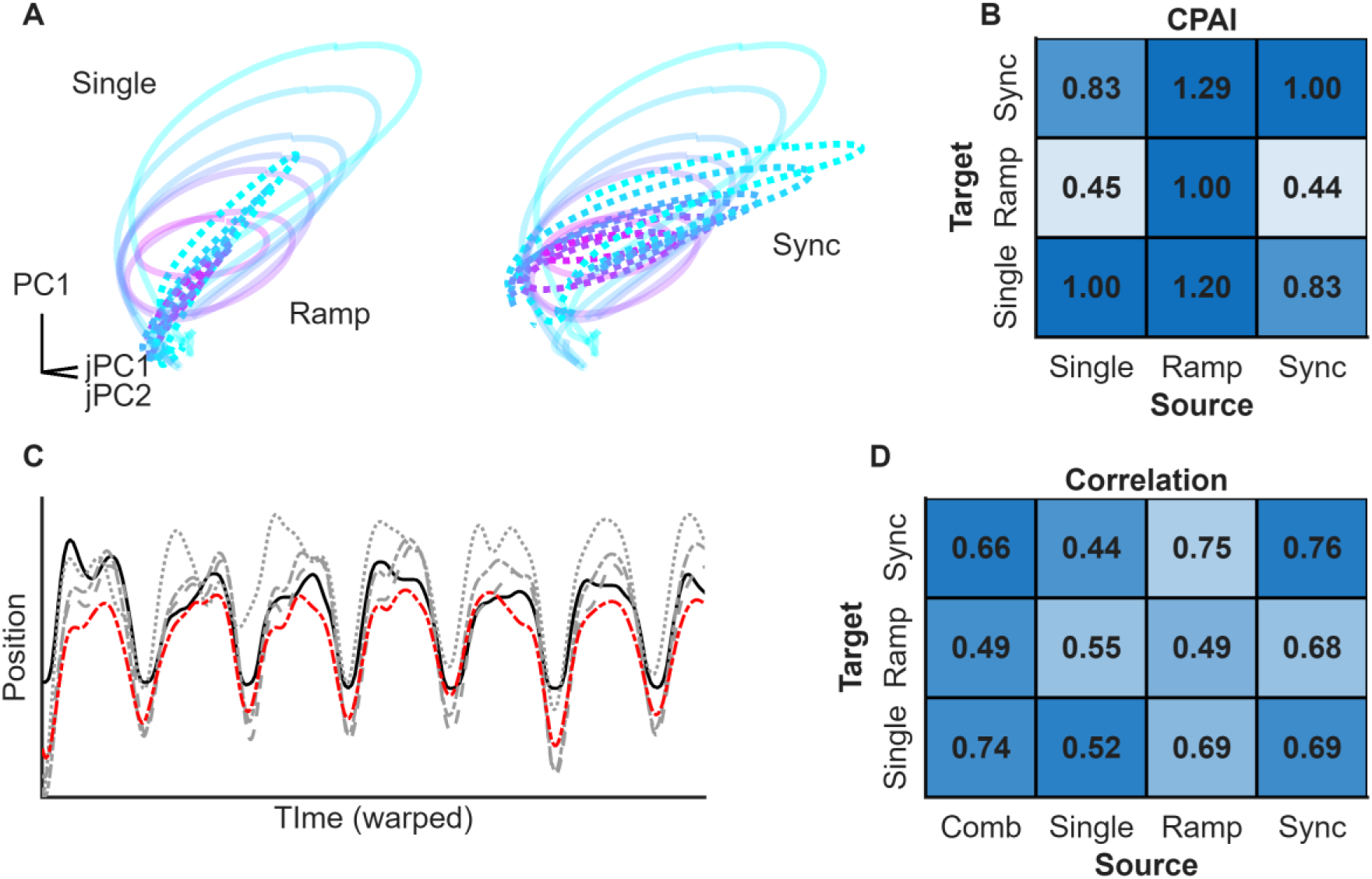
Cross task projections and decoding performance. **A)** Trial and tap-averaged single tempo data projected into its own space (transparent lines) versus when projected into the ramp or syncopated spaces (dotted lines, left and right, respectively). **B)** Cross-projection alignment index for each pair of tasks. **C)** Example cross-task decoding. Black line = ground truth, dashed gray line = single tempo decoder, dotted gray line = ramping tempo decoder, dash-dot gray line = syncopated decoder, dash-dot red line = combined factor space decoder. **D)** Decoder performance (Pearson’s correlation) by source decoder and target task.

Next, we sought to determine if these misalignments impacted decoding performance. We thus trained a linear model on the 5-dimensional space to predict finger position for each task and then evaluated its performance on all other tasks. We found that while correlations were high across tasks (**Figure 6C-D**), the coefficient of determination (R^2^) was especially poor between the ramp task and the other tasks (**Supplementary Figure 9C**), consistent with the cross-projection alignment index. Moreover, this is consistent with the observation that the trajectory radius scaled with kinematic range in the single tempo task but not the ramping task. We reasoned that differences in the tasks may have resulted in differences in the neural activity creating unique spaces for each task separately, but that a common space might exist across tasks. To assess this, we combined all tasks and computed a 5-dimensional space using factor analysis. Repeating the decoding analysis, we found that the combined space performed well across all tasks, though unsurprisingly rarely as well as decoders trained on their own space. Moreover, we used this approach to quantify the dimensionality of the signal (**Supplementary Figure 9D**) and found that decoding performance saturated at approximately 10 dimensions (r = 0.78, R^2^ = 0.61).

## Discussion

In this work we expand upon the existing NHP literature on the representation of rhythm in motor cortex. Primarily, we find that similar rotational dynamics are observed in human motor cortex during rhythmic tapping and that the rotations are modulated by both the tapping range and amplitude within certain contexts. These rotational dynamics have also been observed in motor cortex across a range of behaviors^22,27^ as well as in the spinal cord of locomoting animals^29,30^. Across both fields, a single rotation is observed for each movement suggesting the rotations are a core feature of motor cortex independent of movement type. Indeed, our ability to predict finger position across tasks implies that the relationship between neural activity and kinematics is maintained regardless of the context (**Figure 6D**).

A limitation of this work is that the majority of the data comes from a singular participant. However, many of the results throughout this work are consistent with the results of the single tempo experiment, which we replicated with the second participant. Moreover, the results here are in many ways natural extensions of the NHP literature^17,18^ and did not contradict our hypotheses. Consequently, while not all experiments were replicated, the robustness and consistency of the data imply that they are not limited to the participants we recorded from.

One distinct difference in the results presented here from previous NHP literature^17,18^ is that the stable point appears to be at the apex of the tap cycle instead of the tap event itself (e.g. **Figure 1E**). This is particularly surprising as the tap event coincides with a constant kinematic position across tempos. That is to say that the position of the finger when tapping was constant at that singular point in time but unconstrained at all other time points. Consequently, the fact that the neural trajectory differed most at the tap event conflicts with the notion that the first jPC purely represents position (**Figure 2E,F**), something that is reinforced by the results of the feedback experiment in which the projections along the first jPC are smaller despite the fact that the range of motion is greater (**Figure 3H,I**). Thus, the changes in sensory feedback associated with moving at different tempos – both tactile and proprioceptive – may instead drive these effects. On the other hand, the rhythmic timing literature considers that the tapping times are associated with the subjective beat or pulse that can vary from isochronous to quite complex and weakly periodic^5^. The fact that taps were not aligned in the neural trajectories may reflect a human emphasis for finger control during the dwell period over synchrony across a range of tempos. In contrast, in the case of NHPs, synchrony was of upmost importance given the reward contingencies, potentially producing the convergence of trajectories at tapping times.

The single tempo task also revealed a continuum of kinematic strategies (**Figure 2A**) that indicate different cognitive approaches to rhythmic tapping. At the lowest tempos the kinematics were relatively stable during the elongated dwell period and then punctuated by an abrupt tap and this was reflected in the neural trajectories with an excursion outside of the primary rotational plane(**Figure 2B**). In contrast, at the highest tempos the kinematics were sinusoidal and neural trajectories did not leave the rotational plane. These differences may reflect a shift from a reactive to anticipatory timing mechanism that likely involve different cortical regions, e.g. prefrontal and parietal at the lower tempos versus cerebellum at the higher tempos^31^. Whether this represents a binary shift in timing mechanisms or a spectrum is unclear.

Regarding the role of feedback, our initial hypothesis was that the participant would use the high-salience tactile event to align their taps but would be unable to do so in the no feedback condition. However, we saw extremely robust sinusoidal movements in the kinematics that were as well aligned with the tones as those in the feedback condition. This refutes our initial hypothesis yet revealed a more surprising result in that the magnitude of the dynamics was dependent on the presence of feedback (**Figure 3H,I**). This supports the argument that while tactile feedback may not be strictly required to produce many types of movement it is still integrated into motor cortex, which helps align the neural activity that generates these movements. Additionally, this provides more evidence that sensory signals are integrated into motor cortex consistent with recent literature^32–34^.

One surprising result was the lack of modulation during the preparation phase of the tempo preparation task (**Figure 4B,C**) given that previous NHP literature has reported this^35^. Recent research however, has indicated that this may be a consequence of the association of the reward with progression through the trial and consequently related to the training^36^, something that we did not need to do for our experiments. Alternatively, the lack of activity in the preparation phase could be attributed to the specific instructions given. We explicitly told C1 not to imagine tapping, but instead to observe the tempo and prepare to join it when cued. If he were cued to imagine tapping at the appropriate tempo the opposite may have occurred. Given that previous research has shown that many regions are involved in rhythmic activities^13,19^, it stands to reason that signatures of tempo preparation may appear there while primary and pre-motor cortex are restricted to kinematic representations in the absence of reward driven effects. With regards to the continuation phase of the task, the lack of behavioral variability – and thus neural variability - precluded extra analysis. Given the results of the single tempo experiment, we would expect that if drift was observed, the trajectories would follow the kinematic and tempo dependence observed in the single tempo and ramp experiments, though they could exist in a subspace unique to imagined movements^37^.

Both the single tempo and mixed tempo experiments raise interesting questions about how we both know when to tap and how we know when to change the tempo at which we are tapping. Low dimensional projections at the slower tempos in the single tempo experiment and the primary tempo in the mixed tempo experiment revealed consistent excursions orthogonal to the rotational plane which we interpret as a wait signal as it corresponds with the dwell period (**Figure 2E**). While our inability to predict tempo extremely well across conditions could be attributed to trial-specific effects, the fact that we could not perfectly disentangle primary and secondary tempos during the dwell period (**Figure 5L**) implies that other brain regions such as the supplementary motor area of prefrontal cortex^19^ are instead responsible for keeping track of rhythm in the absence of movement as well as transitions in tempo.

## Methods

### Participants

This study was conducted under an Investigational Device Exemption from the U.S. Food and Drug Administration and approved by the Institutional Review Boards at the Universities of Pittsburgh and Chicago. The clinical trial is registered at ClinicalTrials.gov (NCT01894802). Informed consent was obtained before any study procedures were conducted. Participant C1 (male, m), 55-60 years old at the time of implant, presented with a C4-level ASIA D spinal cord injury (SCI) that occurred 35 years prior to implant. C1 retains partial function of the contralateral arm and wrist but can only achieve hand movements with tenodesis grip. Participant C2 (male), 60-65 years old at time of implant, presented with a C4-level ASIA D spinal cord injury (SCI), along with a right brachial plexopathy, that occurred 4 years prior. C2 retains limited shoulder and wrist function.

### Cortical implants

We implanted four microelectrode arrays (Blackrock Neurotech, Salt Lake City, UT, USA) in each participant. The two arrays (one medial and one lateral array) in Brodmann’s area 1 of S1 were 2.4 mm x 4 mm with sixty 1.5-mm long electrode shanks wired in a checkerboard pattern such that 32 electrodes could be recorded from or stimulated. The two arrays in motor cortex were 4 mm x 4 mm with one-hundred 1.5-mm long electrode shanks wired such that 96 electrodes could be used to monitor neural activity. The inactive shanks were located at the corners of these arrays. Two percutaneous connectors, each connected to one sensory array and one motor array, were fixed to the participant’s skull. We targeted array placement during surgery based on functional neuroimaging (fMRI) or magnetoencephalography (MEG) of the participants attempting to make movements of the hand and, within the constraints of anatomical features such as blood vessels and cortical topography – see ^38^ for a full description of the implantation approach.

### Task instructions

Each participant was instructed to tap a metal plate (see *Tap synchronization*) using their index finger on each metronome tick (when present). Participants were encouraged to complete a full trial (30 seconds to 3.5 minutes) and were allowed unlimited breaks in-between trials as requested to minimize fatigue. Due to the large number of taps that each participant completed, we worked with them to position their forearms and wrist so that they could sit comfortably for extended periods of time.

### Audio cue delivery

Audio files were constructed using Matlab (2024a, Mathworks, MI, USA). Briefly, a metronome tick sampled at 24 kHz (see Supplementary Files) was repeated with the appropriate temporal spacing to produce the audio file for a given trial. Before each trial, a bell sound was played to indicate to the participant that the trial would begin in 3 seconds. Audio was played through Fostex RM-3 rack mount speaker system (Foster Electric Co., Tokyo, Japan) and the reference/headphone-out port was connected to the neural acquisition system with a 1/4” inch to BNC connector which was sampled at 10 kHz. Audio files can be found in the *Supplementary Materials* while the code used for creating the audio files can be found in the code repository.

### Single tempo tasks

To measure the ‘natural’ tempo of each participant, we also asked them to tap at either a “comfortable”, “slow”, or “fast” tempo. We recorded this data for approximately 30 seconds to determine the bpm of each natural tempo.

For the simple tempo task, on each trial participants were played 7 audio files with metronome sounds distributed at 30 to 210 beats per minute (BPM). Each of the 7 files was 30 seconds in duration and were played in a random order within a given block. 5-10 blocks were recorded for each participant.

To investigate the participant’s ability to maintain a tempo before and after entrainment, we added a visual cue to the simple task where a green fixation cross indicated they were to tap while a white fixation cross indicated not to. In this task we selected 3 tempos between 30 and 150 bpm and each audio file was 30 seconds long. On half of the trials the green fixation cross appeared immediately while on the other half the white fixation cross was displayed for 10 seconds before the green fixation cross appeared for the latter 20 seconds of the audio file. After the audio file finished playing, the green cross remained, and the participants were encouraged to continue tapping at the appropriate tempo for a further 10 seconds.

Contact of the finger with the tap plate represents a discrete and abrupt event that the participants intuitively attempted to align with each metronome tick. To determine if sensory feedback and rapid deceleration influenced the participants, we interleaved trials in which the participants were instructed to perform the simple tempo task either while contacting the tap plate or ‘tapping the air’. We again selected 3 tempos between 30 and 150 bpm and played each audio file for 30 seconds. As this task required that the participants not contact the receiving plate on some trials, we marginally elevated their wrists so that they could comfortably perform similar ranges of motion without contacting the tap plate.

### Ramp task

In the interest of maximizing the number of unique tempos and examining what happens when the target tempo slowly drifts, we constructed audio files in which the bpm was either continuously ascending or descending. In both cases there was a lead in period of 10 seconds to enable the participant to entrain before the tempo began changing. Then, each trial was composed of 300 taps where the inter-tap-interval was linearly spaced between 30 and 210 bpm. Consequently, the ramp duration was 195 seconds in duration and the total trial time was 205 seconds. We repeated both the ascending and descending ramps 3 times each.

### Mixed tempo task

All of the previously described tasks consisted of trials in which the tempo was either static or monotonically changing. As musical beats are often composed of rhythms, we also constructed audio files in which the bpm of the taps varied according to integer ratios. First, we chose a primary frequency between 30 and 90 bpm followed by a secondary frequency that was between double the base frequency and 180 bpm. We then produced ‘bursts’ of taps at the secondary frequency which were spaced by the primary frequency. Second, we chose a base frequency between 60 and 120 bpm and secondary frequencies between 30 bpm greater than the base frequency and up to 240 bpm. Then, we alternated taps at the primary and secondary frequency so that a constant ratio of tap times was produced.

### Tap synchronization

To precisely measure contact and release time of the finger at a temporal resolution greater than image capture allows, we used a custom current flow detector. The 5v+ output of an Arduino board (Uno Rev3, Arduino LLC, MA, USA) was split with one output connected to a 20 MOhm resistor which then fed to ground while the other lead contacted the participant’s finger. A metal tap plate was then also attached to the ground line on the Arduino such that when the participant tapped the tap plate, they closed the 5v circuit. This then triggered the digital output port of the Arduino to change from high to low. This port was connected to the neural acquisition system such that the contact status of the finger could be sampled at 1 kHz. To influence from the 5v Arduino line upon the neural data, each participant’s finger was insulated with a latex glove.

### Kinematic tracking

We recorded finger, hand, and wrist kinematics with a single camera (Blackfly 3, Teledyne FLIR, OR, USA) pointed towards the hand such that we could determine kinematics in addition to the precise tap timing. Images were cropped (400 x 800 pixels) and captured at 50 Hz in grayscale. The 3v line-out of the camera was sent to a digital input of the acquisition system such that each image could be aligned with the neural data. For automated labelling and tracking of the frames we used JARVIS^39^ to perform keypoint detection of the tip of the index finger, the metacarpal joint, the hand center, and the wrist. Keypoints were then filtered to remove outliers followed by a median filter and gaussian smoothing filter. For the feedback task kinematic peaks were also identified using the *findpeaks* function in Matlab.

### Data acquisition

Neuroplex E headstages (Blackrock Neurotech, UT, USA) were used to connect each pedestal to a NeuroPort system (Blackrock Neurotech, UT, USA). Voltages were sampled at 30 kHz and high-pass filtered (750 Hz) with a 1^st^ order Chebychev filter. An RMS threshold of −4.5 was used to detect spikes from the continuous signal. The audio output was sampled at 10 kHz, the tap line at 1 kHz, and the digital line from the camera at 30 kHz.

### Neural data processing

As each tap had a unique duration, to better compute metrics across taps, neural data between tap events was resampled and spline interpolated such that each inter-tap interval had 200 discrete values. For the *natural tempo* dataset, warped data was smoothed across 3 taps as multiple trials were not collected due to the implicit variability with intrinsically generated tempos. For all other datasets, taps were aligned to audio cues and were identified as valid or invalid based on whether two successive taps were within 15% of the target cue (where the 15% threshold is computed based on the target inter-tap-interval for each pair of taps). Taps were then averaged either by position in the trial (i.e. the 5^th^ tap of each trial) or with a sliding window (i.e. the 1^st^-3^rd^ taps).

### Dimensionality reduction

Natural tempo experiment: smoothed firing rates from each tempo and PCA was performed on all warped taps simultaneously to estimate the top 3 PCs that explained the most variance. Additionally, when jPCA was used^22^, each warped tap was treated as an individual condition.

Single tempo experiment: jPCA was performed on the tempo-averaged tap data (n=7). The jPCA normalization steps were then applied to the raw data and the single-trial data was projected onto the first jPCA plane. The resulting values were subtracted from the normalized data to produce the jPCA subtracted activity – residuals. PCA was then performed on this residual activity to estimate the residual dimensionality. To ensure the identified dimension used was orthogonal to each jPC, the selected PC was the first in which the absolute Pearson correlation between the PCA coefficients and the jPCA coefficients was less than 0.1.

Ramping tempo experiment: jPCA and PCA were used as described above to identify the maximally rotational plane and the residual PCA space. Multiple linear regression was then used on the PC scores of the first 10 dimensions to predict the target tempo of each tap. The regression weights were then normalized by the sum of the absolute values.

Feedback experiment: For jPCA processing, trials were aligned to the tones instead of the physical taps. This alignment difference does not alter the ability of jPCA to find the optimal plane as the covariance matrix is independent of the temporal alignment across trials. Cross projection was achieved by applying the jPCA normalization steps for each dataset before multiplying by the target coefficients. The relative variance was then computed as the ratio of the total variance after projection onto each plane, representing half of the cross-projection similarity index.

Imagination and continuation experiment: To compute the respective jPCA subspaces, neural data was sampled either with tap-aligned neural data (executed subspace) or tone-aligned data (preparation and combined subspaces). Cross-projection of different tap types was accomplished as described above.

Mixed tempo experiment: Each trial type was composed of a repeating beat sequence of 2-6 audio cues. For each trial type, averaging was first performed across trials and then all sequences within the trial were averaged. Sequence tap events were then flattened before jPCA and PCA were applied as described in the single tempo experiment.

### Trajectory metrics

To compute the radius of each trajectory, it was essential to minimize biasing of non-uniform phase distributions. Consequently, tap data was projected onto the first jPCA plane (jPC1,2), then, the phase of each point was computed and binned into 16-equal segments. The average of each segment was computed, mean centered, and then the mean distance from the center computed as following:

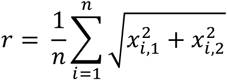

Where *x* is the 2-dimensional matrix of binned jPCA scores in which rows correspond to time points and columns correspond to dimensions. For the mixed tempo experiment, trajectories were instead divided into half-rotations to account for the transition periods between tempos.

Similarly, to compute the length of each tap in neural space, the norm of the difference in the projections along the time dimension was computed:

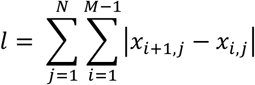

Phase, angle, and rotational strength were all computed as in Churchland et al. 2012^22^.

Tangling was computed in a modified version of that described in Russo et al. 2018^27^. Data was projected into the jPCA + PCA space described above and tangling was computed in those dimensions instead of in raw PCA space. Tangling was computed as the maximum ratio of the difference in derivatives over the distance for all pairs of points within each condition:

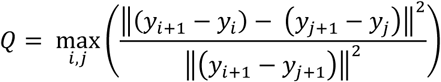

Where *y* represents the neural data projected into N-dimensional space.

## Data Availability

Data will be made available in an online repository upon publication.

## Data and code availability

All data is stored at the Data Archive BRAIN Initiative (https://dabi.loni.usc.edu/dsi/XXXX) and code for analysis is available on GitHub (https://github.com/CorticalBionics/Groove).

## ACKNOWLEDGMENTS

This paper is dedicated to Sliman Bensmaia, may his passion for music and neuroscience inspire us all. We would also like to thank the participants for their generous contribution to the advancement of science as well as members of the Cortical Bionics Research Group for their valuable input on this project. This work was supported by the National Institute of Neurological Disorders and Stroke of the National Institutes of Health under Award Numbers, T32 NS121763, UH3 NS107714, R35 NS122333, and R01 NS130302.

## Extended Figures

**Supplementary Figure 1.**
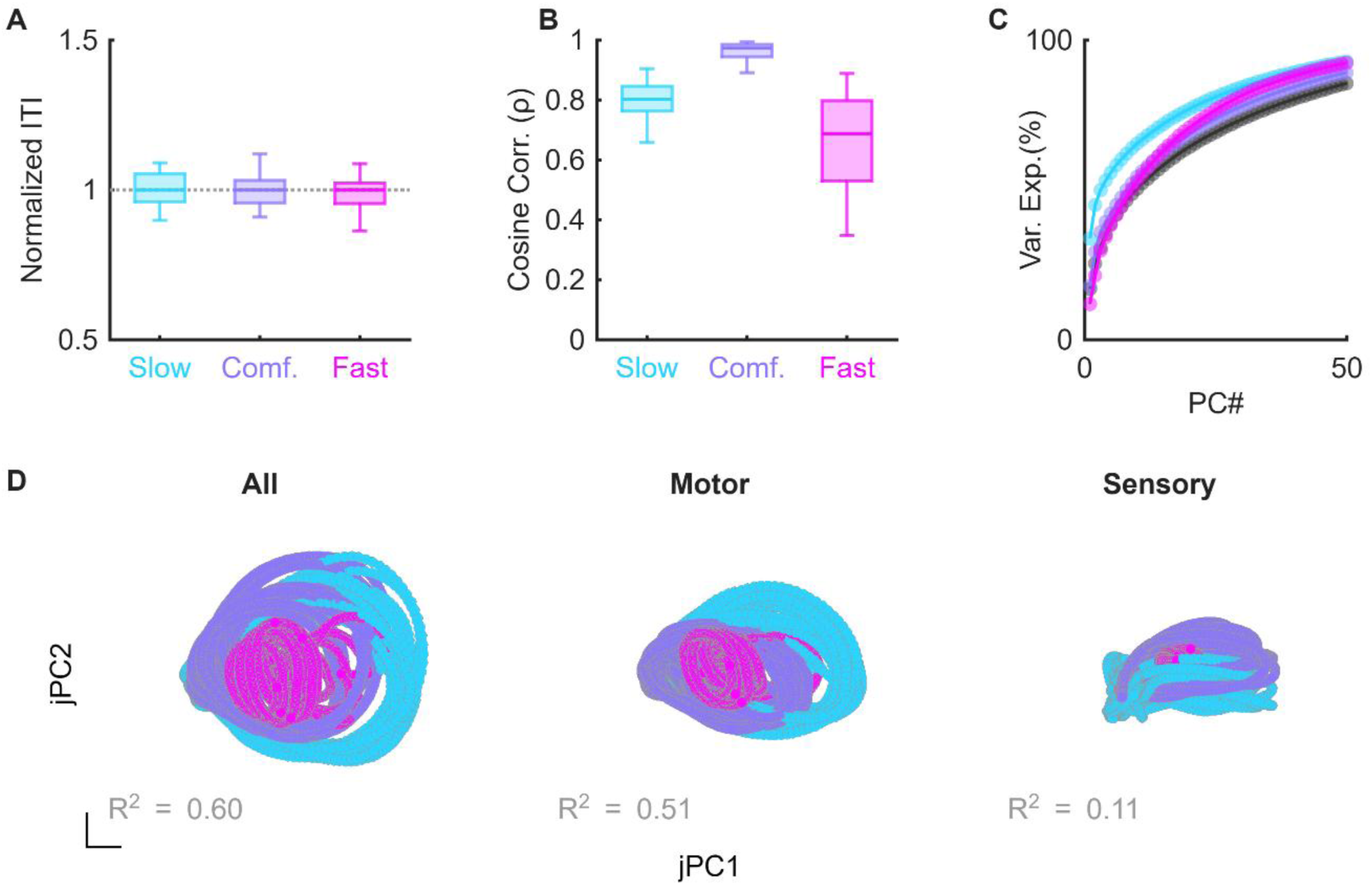
Kinematic and neural properties during natural tempos. **A)** Inter-tap intervals relative to the median inter-tap interval. **B)** Distribution of correlations for each tap with a cosine curve. **C)** Cumulative variance explained by successive PCS for individual tempos and all tempos combined (black). **D)** jPCA projections when computed on all electrodes, motor electrodes, or sensory electrodes. Data from C1.

**Supplementary Figure 2.**
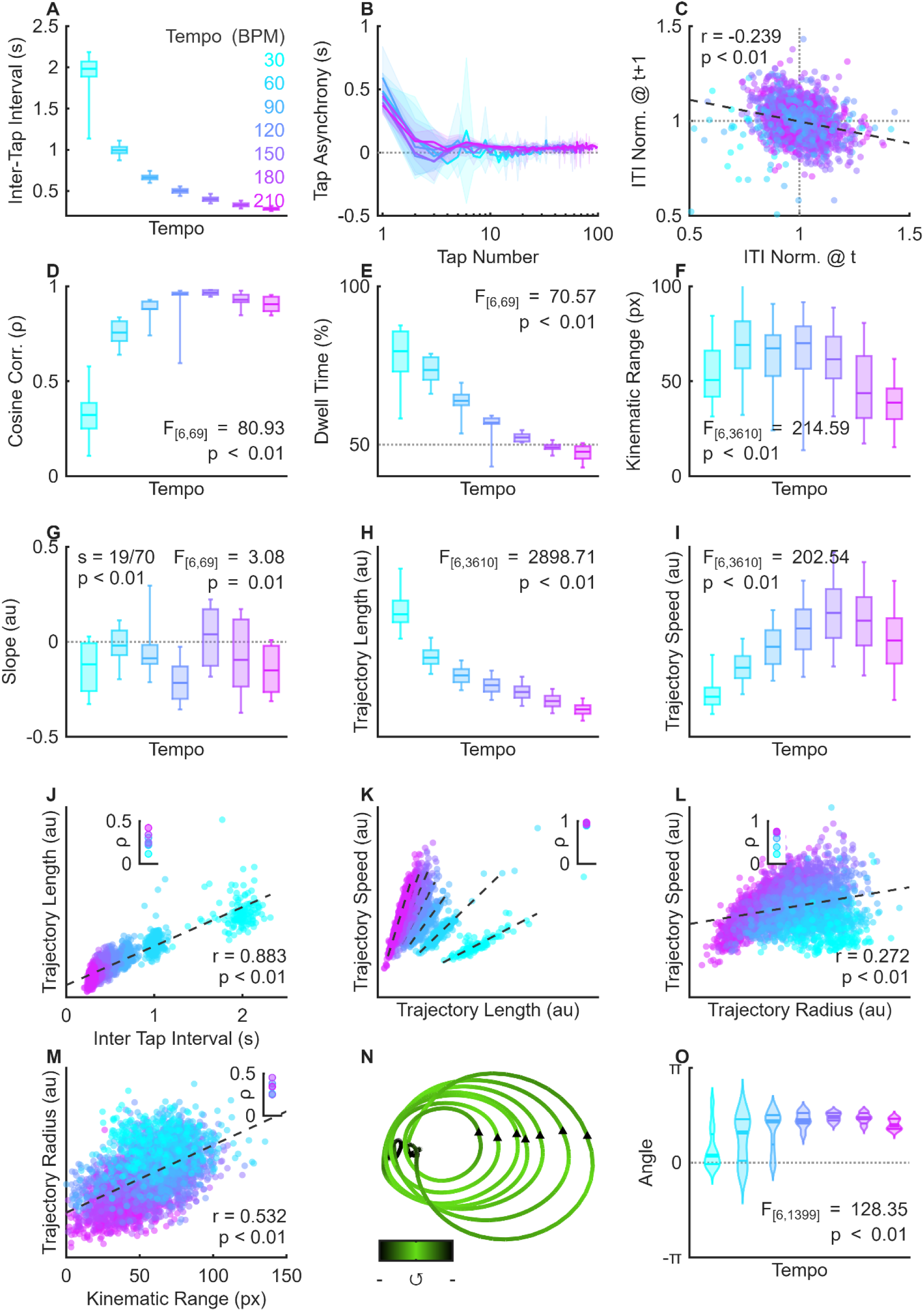
Kinematic and neural properties during single tempo tapping in Participant C1. **A)** Absolute inter-tap interval for each of the cued tempos. Box and whiskers show the 5^th^, 25^th^, 50^th^, 75^th^, and 95^th^ percentiles. **B)** Latency between each auditory cue and the nearest tap event. Positive values reflect taps that occurred after the cue while negative values reflect those that preceded the cue. **C)** Pearson correlation between inter-tap intervals for each tap and that of the subsequent tap. **D)** Correlations between a negative cosine wave and the kinematics for each tap across tempos. **E)** Percent of each tap spent in the dwell phase (top 50% of the kinematic range) for each tempo. **F)** Distribution of kinematic ranges for each tempo. **G)**Slope in PC space of centroid distance from the stable point over the first 5 taps. **H)** Trajectory length and **I)** speed across 3-dimensions for each tempo. **J)** Relationship between inter-tap interval and the length of the neural trajectory for each tap. Dashed line indicates Pearson correlation across tempos while the inset shows correlation within tempo. **K)** Relationship between trajectory speed and length. Dashed lines indicate best fit line within each tempo while the inset indicated correlation values for each fit. **L)** Relationship between trajectory radius and trajectory speed. Dashed line indicates Pearson correlation across tempos while the inset shows correlation within tempo. **M)** Relationship between kinematic range and trajectory radius. Dashed line indicates Pearson correlation across tempos while the inset shows correlation within tempo. **N)** jPC1,2 view of tap-averaged trajectories colored by angle. **O)** Distribution of rotation angles for each tempo. Thick line shows median while thinner lines show 25^th^ and 75^th^ percentiles.

**Supplementary Figure 3.**
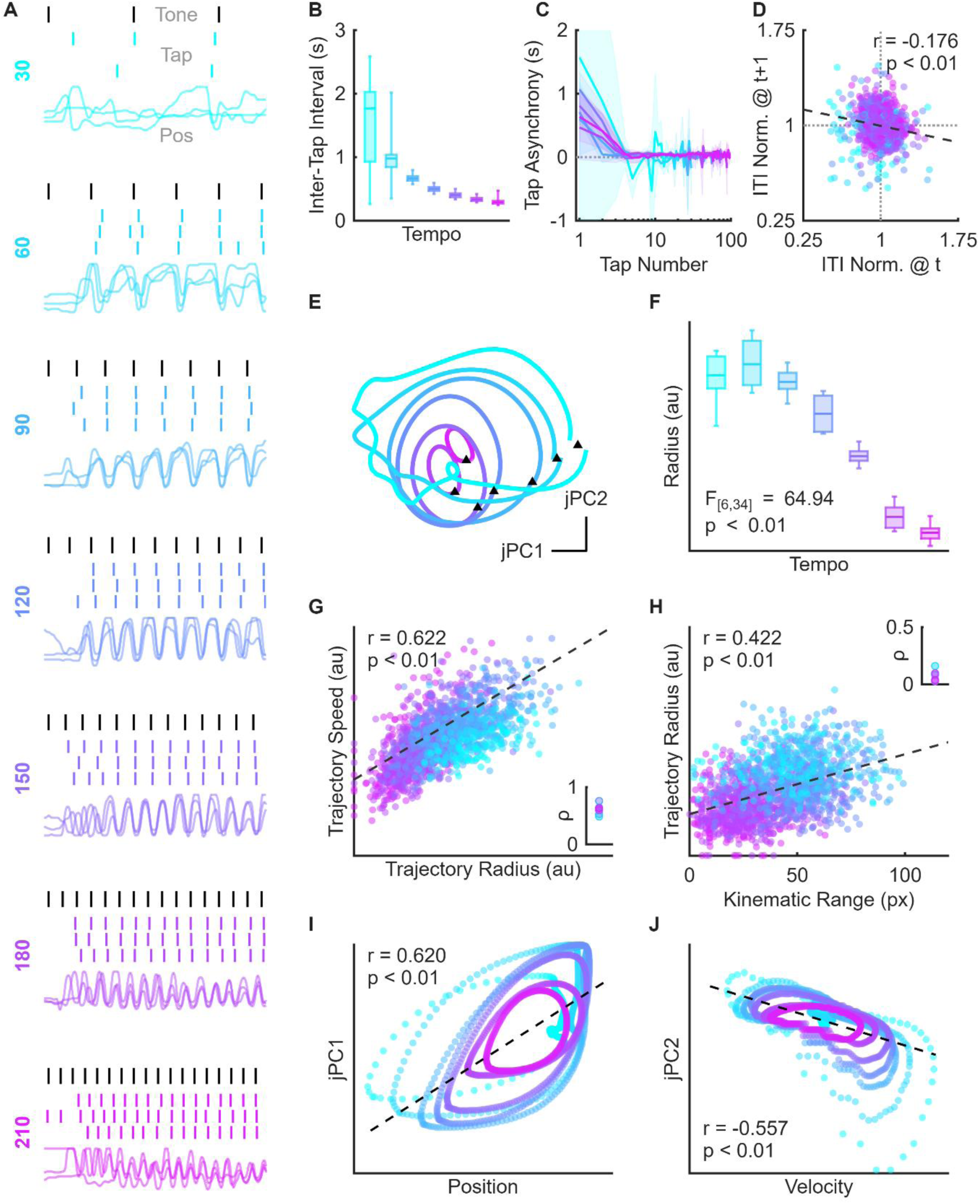
Single tempo analysis in participant C2. **A)** Example kinematics from 3 trials. Black ticks indicate auditory cue times, colored ticks indicate the time at which the participant contacted the capacitive sensor, and colored traces indicated the vertical position of the fingertip over time. **B)** Absolute inter-tap interval for each of the cued tempos. **C)** Latency between each auditory cue and the nearest tap event. **D)** Pearson correlation between inter-tap intervals for each tap and that of the subsequent tap. **E)** Rotational plane view of the tap-averaged data colored by tempo. **F)** Distribution of neural radii for each tempo. **G)** Relationship between trajectory radius and trajectory speed. Dashed line indicates Pearson correlation across tempos while the inset shows correlation within tempo. **H)** Relationship between kinematic range and trajectory radius. Dashed line indicates Pearson correlation across tempos while the inset shows correlation within tempo. **I)** Relationship between tap-averaged position and jPC1 score for each tempo. **J)** Relationship between tap-averaged velocity and jPC2 score for each tempo.

**Supplementary Figure 4.**
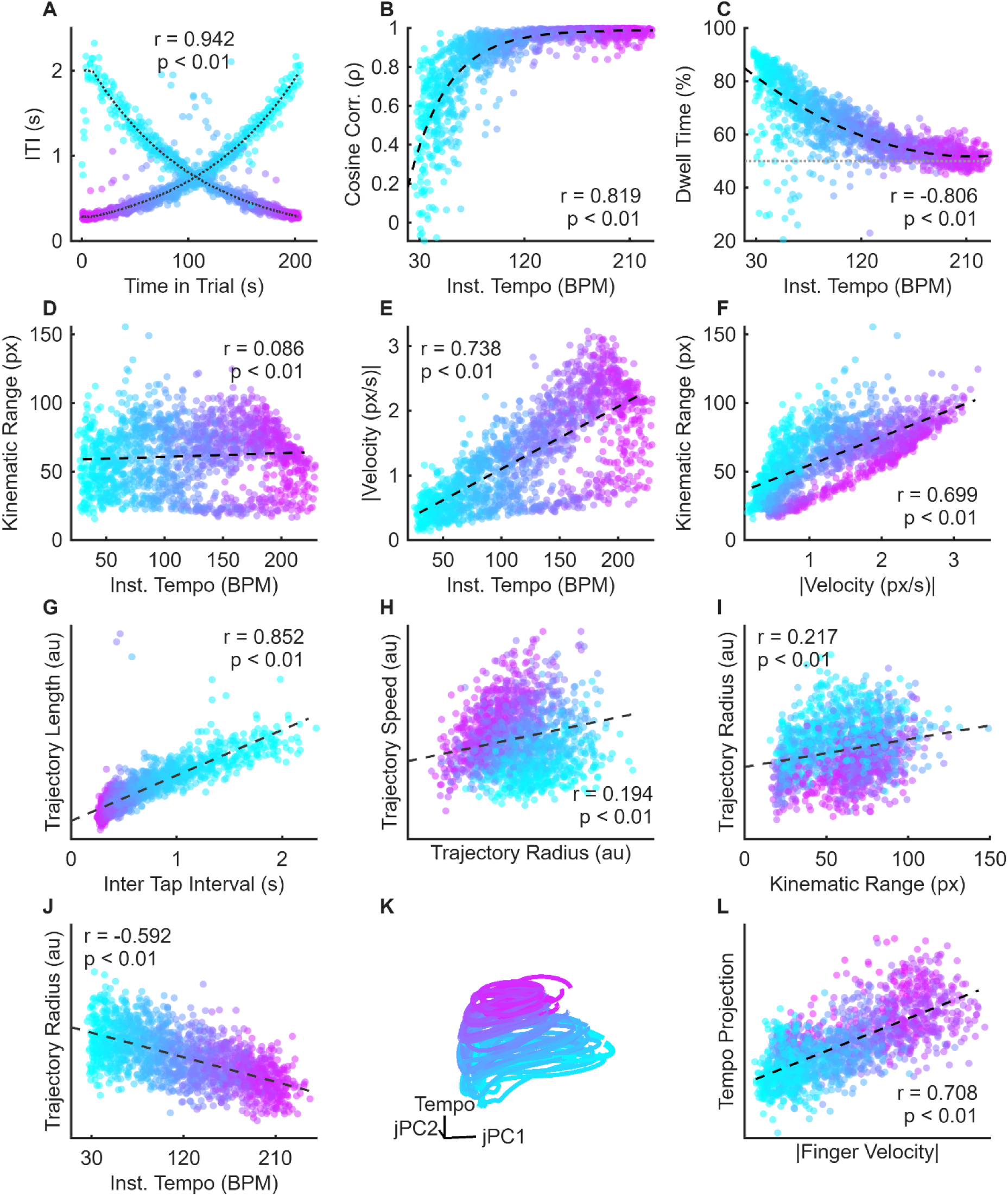
Relationships between neural activity and kinematics during the ramping task. **A)** Absolute ITI for each trial type. Dotted line indicates target ITI for ascending and descending ramps, respectively. Each marker represents a tap event and is colored by the instantaneous tempo. Spearman correlation was computed as the correlation between the target ITI (dotted line) and observed ITI across trials. **B)** Relationship between instantaneous tempo and cosine correlation, **C)** dwell time, **D)** kinematic range, and **E)** absolute finger velocity. Dashed line shows best fit. **F)** Relationship between absolute finger velocity and kinematic range. **G)** Relationship between inter tap interval and trajectory length. **H)** Relationship between trajectory radius and trajectory speed, **I)** kinematic range, and **J)** instantaneous tempo. **K)** Descending trial data projected into the low-dimensional space. **L)** Relationship between kinematic range and trajectory radius across all taps. Data from C1

**Supplementary Figure 5.**
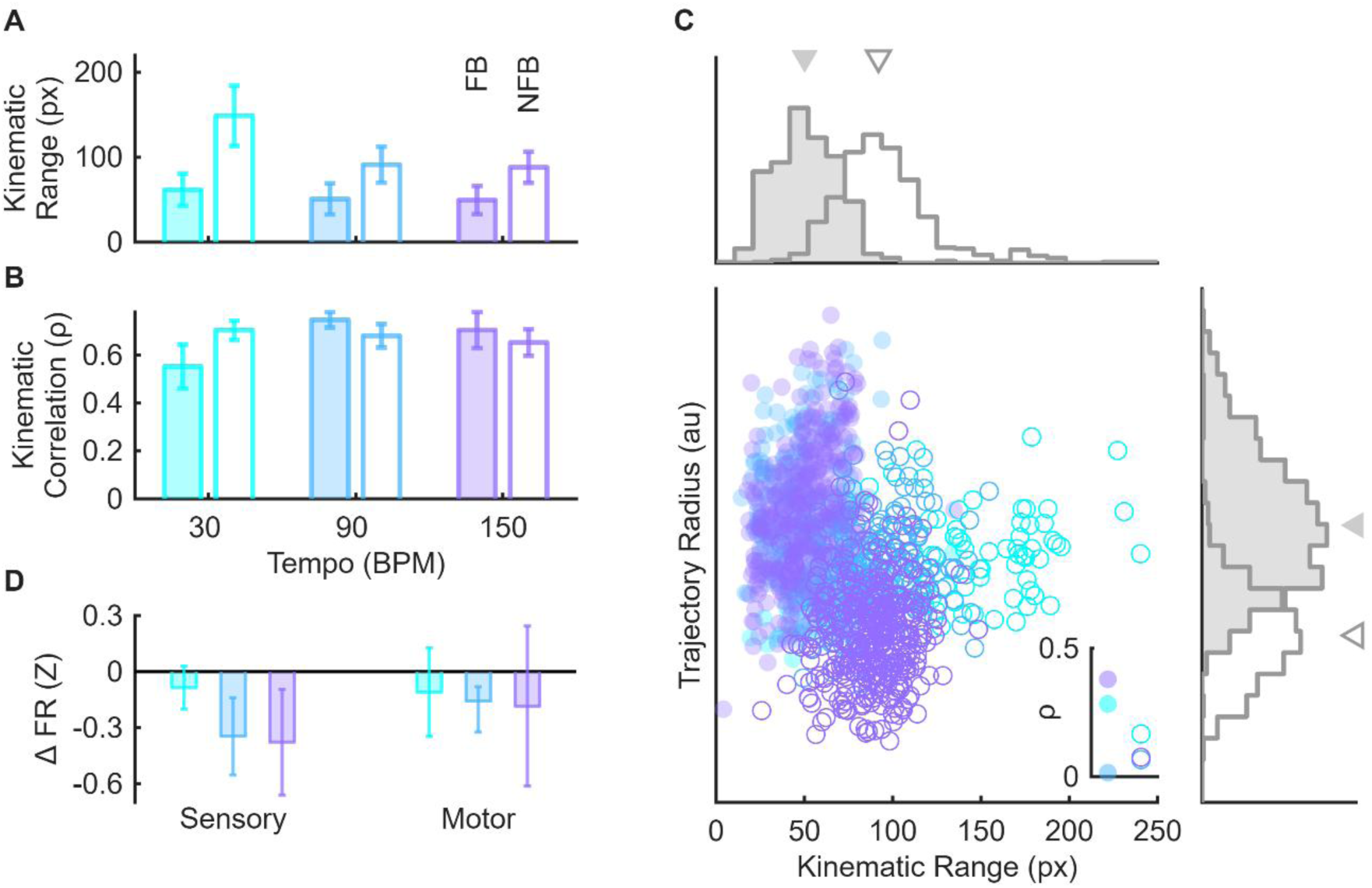
Neural activity and kinematics in the presence or absence of tactile feedback. **A)** Kinematic range (95^th^ – 5^th^) percentile for each tempo and trial type. Shaded bars represent Feedback (FB) trials and empty bars represent No Feedback (NFB) trials. **B)** Within trial kinematic correlations for each tempo and trial type. **C)** Kinematic range and trajectory radius for each trial. Filled markers represent FB taps and empty markers represent NFB taps. **Inset)** Within tempo correlations for each tempo and trial type. **Top marginal histogram)** Distribution of kinematic ranges for FB (filled) and NFB (empty) taps. Arrows above indicate median value for each distribution. **Right marginal histogram)** Distribution of trajectory radii for each trial type Formatting as in the top marginal histogram. Data from C1.

**Supplementary Figure 6.**
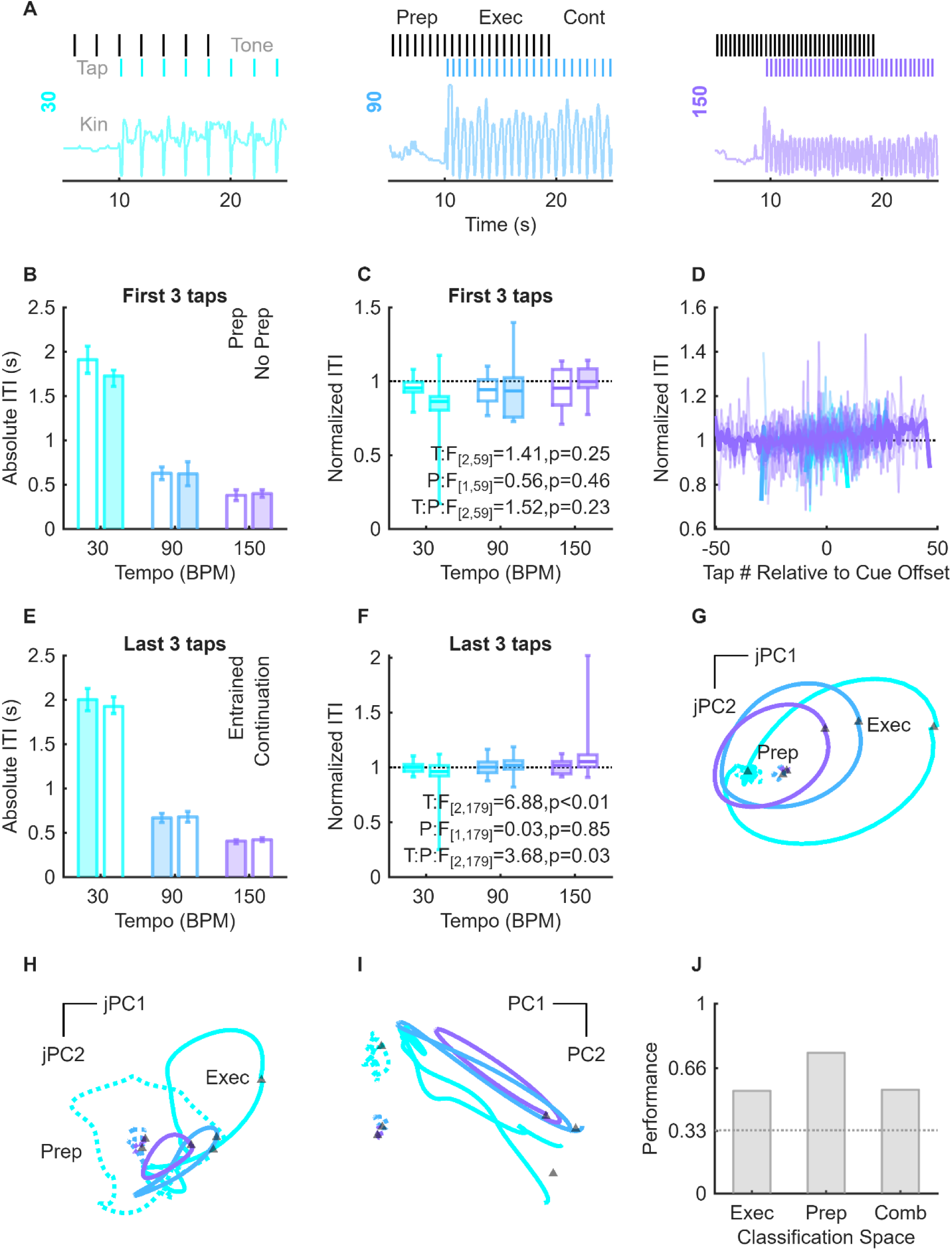
Behavioral metrics with and without auditory cues. **A)** Tones, taps, and kinematics for an example trial at each tempo. Trials consisted of phases with tones but no tapping, tones and tapping, and tapping without tones. **B)** Absolute and **C)** normalized inter-tap intervals for the first 3 taps split by trials with and without preparation time. **D)** All inter-tap intervals normalized to the target tempo relative to cue onset. Individual trials shown in lighter shades, trial average shown with the darker shade. **E)** Absolute and **F)** normalized inter-tap intervals for the last 3 taps during the entrained period (when cues were present) and continuation period (when cues were absent). **G)** Trial averaged trajectories for preparation and physical taps projected into the jPCA plane fit to the physical tap data. Solid lines indicate physical taps, dotted lines preparation period, and triangular markers tap events. **H)** Trial-averaged physical and preparation trajectories projected into the top jPCA plane fit to the preparation period data. **I)** Trial averaged trajectories instead projected onto the top 2 principal components computed on both the preparation and executed periods. **J)** Classification performance of the prepared tempo using LDA on PCA scores when projected into 3D subspaces computed on executed, prepared, or combined datasets. Dotted line indicates chance. Data from C1.

**Supplementary Figure 7.**
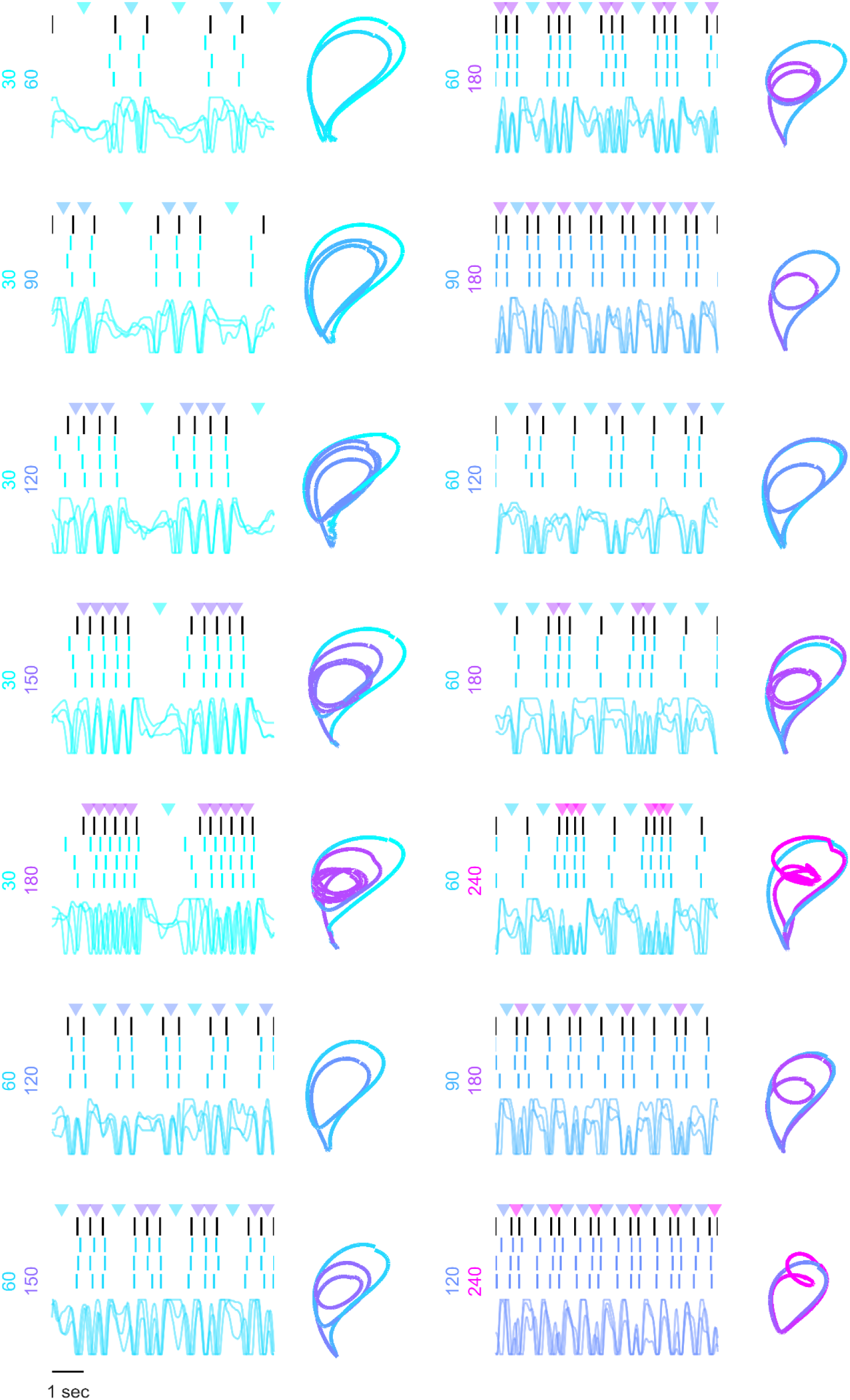
Mixed tempo kinematics. Example trials for each of the 14 conditions. Primary and secondary tempo shown on as y-labels for each. Sequence averages trajectories shown for each.

**Supplementary Figure 8.**
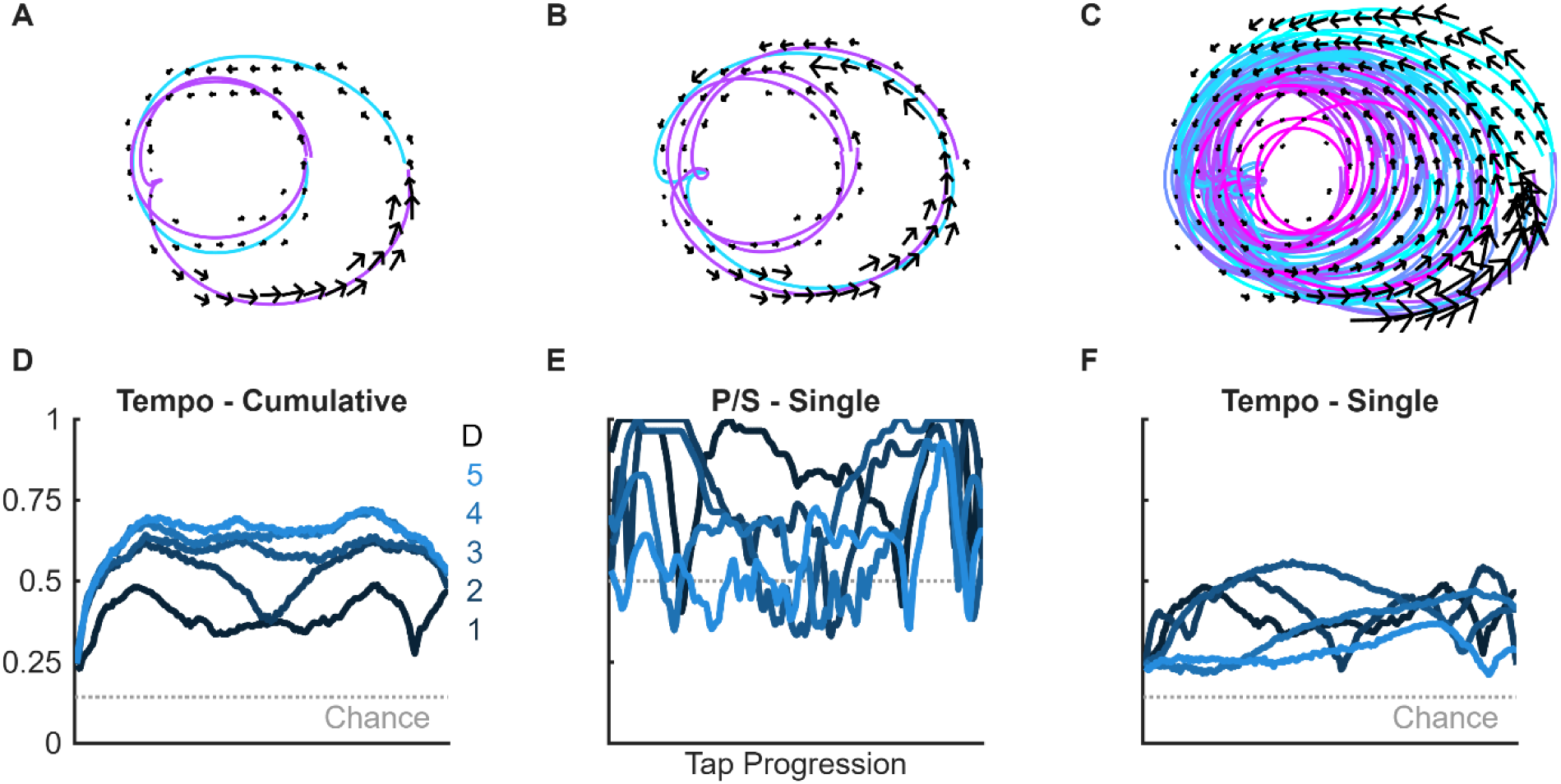
Tangling of mixed tempo trajectories. **A-C)** Larger version of insets from Figure 5D-F with overlayed with arrows, the size of which indicates the vector strength at each location. **D)** Tempo classification with increasing number of cumulative dimensions. **E)** Single dimension classification performance within or **F)** across conditions. Data from C1.

**Supplementary Figure 9.**
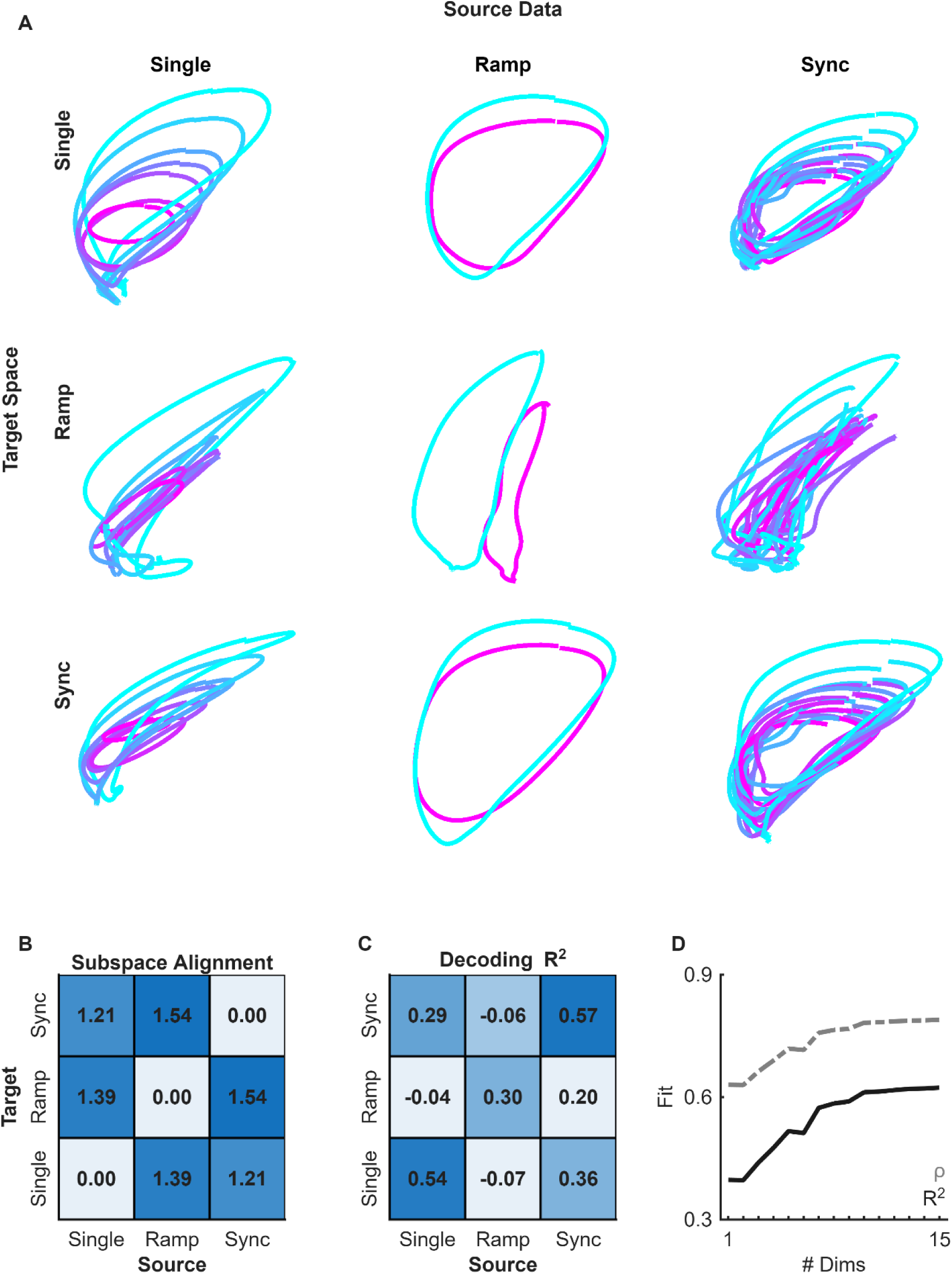
Scaled cross-task neural projections and decoding. **A)** Condition tap-averaged trajectories of each task projected into each task space. In the case of the ramp task, the ascending trajectory is in blue while the descending trajectory is in magenta. Note that for the syncopated task these are not sequence-averaged trajectories. **B)** Subspace alignment of the 5-dimensional space between tasks. **C)** Cross-task decoding performance as measured with residual variance. **D)** Decoding metrics for the cross-task factor analysis linear decoder as a function of the number of dimensions.

